# The lingering legacy: Resilience as a pathway linking retrospective organisational support and police retirement adjustment

**DOI:** 10.64898/2026.04.08.26349526

**Authors:** Eleftheria Vaportzis, Warren Edwards

**Author notes:** Corresponding author details: Full name Eleftheria Vaportzis, Postal address Department of Psychology, University of Bradford, Bradford, BD7 1DP, UK.

## Abstract

This study evaluated retirement adjustment in UK retired police officers (N = 289) across two analytical models. The moderation model revealed a non-significant overall interaction, though exploratory Johnson–Neyman analysis identified an emergent region at 32.07 years post-retirement, where perceived organisational support significantly predicted adjustment. This suggests a long-term organisational legacy effect, wherein retrospective perceptions of service value serve as a durable psychological resource in later life. Conversely, the mediation model confirmed psychological resilience as a significant statistical mediator directly linking support to adjustment. The findings support lifelong post-service wellbeing strategies, as organisational relationships influence adjustment decades after retirement.

---

Policing is recognised as a high-demand occupation characterised by sustained exposure to stressors (Violanti et al., 2017). Empirical research indicates that police officers operate under conditions of chronic operational and organisational strain that significantly affect psychological wellbeing over time (Andersen et al., 2023). These stressors accumulate across officers’ careers and extend into retirement, exhibiting strong associations with long-term psychological outcomes (Lennie et al., 2025). Leaving policing is a complex transition involving shifts in identity, routine, and social structure, all of which may influence retirement wellbeing (Porter and Lee, 2024).

Retirement transition is particularly salient in policing due to the strength of the police identity (Rowe et al., 2023). This identity is shaped throughout officers’ careers which are often marked by repeated exposure to traumatic events, increasing the risks of long-term psychological difficulties (Birch, 2026). While retirement is theoretically conceptualised as a dynamic, resource-based transition process within general population models (Wang, 2007; Wang et al., 2011), a central question remains: in what ways do the post-service adjustment trajectories of police officers diverge from these standard patterns? Unlike typical civilian transitions, policing is characterised by extreme structural demands and an intense occupational identity that can cast a long shadow over post-service life (Janssens et al., 2021), sometimes manifesting in severe psychological crises and transition difficulties long after leaving active duty (Violanti, 2026). Examining these transitions helps clarify how high-strain occupational contexts shape retirement trajectories differently from general population norms.

Addressing this gap is important not only for extending theoretical models of retirement but also for advancing understanding of how extreme occupational contexts reshape established adjustment trajectories. This necessity is mirrored by recent UK policy developments which frame officers’ wellbeing as a policy priority and a moral obligation (Home Office, 2024). Therefore, understanding how officers adjust after leaving the service is essential.

Past research on police wellbeing has disproportionately focused on serving officers with limited work on the wellbeing of retired officers. Quantitative studies indicate that serving and retired officers experience elevated rates of chronic occupational stress and psychological distress compared to the general population (Kyranides et al., 2025; Violanti et al., 2017). Qualitative studies further illuminate how these structural pressures manifest subjectively as complex identity transitions, with many retired officers reporting a heavy reliance on personal coping resource (Lennie et al., 2025). Their service experiences and individual psychological resources may play a role in shaping post-retirement adjustment (Birch, 2026). While these factors are recognised, the specific pathways through which career-long trauma and individual resources interact to determine long-term adjustment require further investigation (Porter and Lee, 2024).

According to the Organisational Support Theory, perceived organisational support is defined as the extent to which individuals believe their organisation values their contributions and cares about their wellbeing (Eisenberger et al., 1986). This theory posits that perceived organisational support is governed by the norm of reciprocity; when officers perceive high levels of support, they feel an obligation to care about the organisation’s welfare and reach its goals (Kurtessis et al., 2017). In policing, this reciprocal social exchange is foundational to building psychological attachment and resilience (Oscar Kilo, 2024; Kurtessis et al., 2017). When organisational stressors, such as poor leadership or insufficient support, suggest a breach in this exchange, it results in adverse psychological outcomes and increased intentions to leave policing (Oscar Kilo, 2025; Violanti et al., 2017). Where chronic exposure to trauma strains psychological reserves, institutional backing functions beyond immediate job performance; it sets the baseline for a psychological contract (Rousseau, 1989) that persists long after active duty ceases.

To further integrate this framework with individual coping, we draw upon the multi-system model of resilience (Liu et al., 2017). Within this perspective, perceived organisational support acts not only as a reciprocal exchange mechanism but also as an external, macro-system source of resilience. Rather than operating in a vacuum, structural institutional support interacts with individual psychological assets, such as resilience, understood here as a dynamic, multi-systemic process of adapting to adversity over time to shape how officers cope with strain (Denckla et al., 2020; Southwick et al., 2014).

In addition to organisational influences, individual psychological resources are important to retirement adjustment (Lennie et al., 2025). Resilience has traditionally been conceptualised as a trait reflecting the capacity to recover from stress and adapt to adversity (Smith et al., 2008). However, contemporary psychological literature increasingly defines resilience as a dynamic, multi-systemic process of adapting to adversity over time (Southwick et al., 2014; Denckla et al., 2020). In occupational contexts such as policing, this shift highlights resilience not merely as an inherent fixed asset, but as an evolving capacity shaped by ongoing operational stressors and environmental contexts (Janssens et al., 2021). Although officers may demonstrate higher resilience than the general population (Vaportzis & Edwards, under review), this capacity is limited and may be constrained by organisational environments. Conservation of Resources theory provides a useful framework for understanding these dynamics (Hobfoll, 1989). This theory posits that individuals strive to acquire and maintain resources (e.g., psychological resilience, social support), and that stress arises when these resources are threatened or depleted. Within a resource caravan framework, organisational support acts as a critical resource caravan passageway; an environmental condition that fosters, nourishes, or protects personal resources (such as individual resilience; Hobfoll, 2012). According to the theory’s principles, building personal resources requires the investment of existing resources, which is difficult under conditions of chronic stress. Organisational support provides a crucial environmental buffer that reduces ongoing threat appraisal and shields officers from continuous resource depletion. By establishing psychological safety and structural backing, perceived support frees up cognitive and emotional energy, enabling officers to actively invest in and cultivate personal coping mechanisms, thereby generating and sustaining resilience over time. For retired police officers, whose exit from the service strips away structural routine and occupational identity, enduring memories of organisational care serve as a durable contextual resource that shores up personal resilience, thereby mitigating post-service adjustment difficulties.

These theoretical perspectives suggest that retirement adjustment in policing is likely influenced by a combination of organisational and individual factors, as well as the relationships between them. However, there remains a lack of empirical research examining how perceived organisational support and resilience jointly contribute to retirement adjustment, and specifically whether resilience represents a mechanism through which organisational experiences are translated into post-service outcomes. To fill this gap, the present study aims to investigate the relationships between perceived organisational support, resilience, and retirement adjustment among retired UK police officers. We sought to respond to the following research questions:

1. Is perceived organisational support associated with retirement adjustment among retired police officers?
2. Does resilience independently predict retirement adjustment when accounting for organisational and service-related variables?
3. Does resilience mediate the relationship between perceived organisational support and retirement adjustment?

## Method

### Recruitment

Data for this study were drawn from a broader cross-sectional survey examining multiple dimensions of police officer wellbeing (Vaportzis & Edwards, under review). While the larger project addressed various aspects of post-service life, the current study specifically focuses on the relationship between perceived organisational support, resilience, and retirement adjustment.

A cross-sectional survey was designed following guidelines by Kelley et al. (2003). For the purposes of the present analysis, eligibility was restricted to retired police officers in the UK. The survey was distributed online via Online Surveys (JISC, 2021) between November 2025 and March 2026. Data collection concluded after two consecutive days passed without new completions due to a plateau in responses, aligning with the requirements established by our a priori power analysis. Specifically, while an analysis using G*Power (Faul et al., 2009) indicated that a sample of 76 is required to detect a medium effect (f2 = 0.15) for a hierarchical regression with three tested predictors (α =.05, power =.80) and 550 would be needed to detect a small effect (f2 = 0.02), our final sample of 289 is well-powered to detect small-to-medium effects (f2 = 0.04). To ensure data integrity, duplicate entries were minimised using IP tracking and timestamp checks provided by the platform (Wang et al., 2023).

Participants were recruited through a multi-channel outreach strategy targeting the UK policing community. Recruitment materials were disseminated via social media (e.g., Facebook, X) and community groups. Additionally, the study was promoted through the newsletters, websites, and mailing lists of key stakeholder organisations, including the National Association of Retired Police Officers (NARPO), the Police Superintendents’ Association, and the College of Policing. These organisations were provided with study materials and a direct survey link for dissemination to their members. Snowball sampling was also encouraged, with participants invited to share the survey within their professional networks. All recruitment materials explicitly stated the eligibility criteria, ethical approval details, and research team contact information. Ethical approval was granted by the Humanities, Social, and Health Sciences Research Ethics Panel at the University of Bradford (Reference Number E1350). All participants provided informed consent prior to participation.

## Survey design

### Demographic and service questions

Demographic questions included participants’ age, sex assigned at birth, ethnic group, marital status, and current living situation. Service questions included UK service region, highest rank attained, service years, voluntariness of exit, and retirement length. Service years and retirement length were converted from months to decimal years (e.g., 1 month = 0.08) to ensure linear consistency in the models.

### Retirement Adjustment Scale (RAS; Wells et al., 2006)

RAS is a 13-item measure assessing retirement adjustment. Items (e.g., “*I enjoy being retired”*) are rated on a 5-point Likert scale (1 = Strongly disagree – 5 = Strongly agree). To ensure that higher scores consistently reflected better retirement adjustment, the negatively phrased items were reverse-coded. Scores range between 13 and 65. Cronbach’s α was good (.89).

### Brief Resilience Scale (BRS; Smith et al., 2008)

BRS is a 6-item measure designed to assess the ability to recover from stress. Items (e.g., *“I tend to bounce back quickly after hard times”*) are rated on a 5-point Likert scale (1 = Strongly disagree – 5 = Strongly agree). To ensure that higher scores consistently reflected a greater capacity for resilience, the negatively phrased items were reverse-coded. Higher mean scores indicate greater resilience. Cronbach’s α was acceptable (.71).

### Adapted Perceived Organizational Support (Barnes et al., 2013)

This adapted version of the Perceived Organizational Support scale modifies 6 items to reflect military contexts (e.g., *“My Force cared about my wellbeing”*). It measures retrospectively reported organisational support, capturing how former officers evaluate and remember the support provided by their organisation during their active service, rather than real-time ongoing support. Items are rated on a Likert scale (1 = Strongly disagree – 5 = Strongly agree). Scores range between 6 and 30 with higher scores indicating stronger perceived organisational support. Cronbach’s α was excellent (.93).

This scale was chosen because high-reliability measures assessing career-long institutional support tailored specifically for retired emergency services personnel are sparse. Given the structural parallels between military and police institutions, specifically strict hierarchical chains of command, institutional discipline, and exposure to chronic operational trauma, this adapted scale provides a psychometrically sound measure of institutional support within uniformed occupations (Barnes et al., 2013; Violanti et al., 2017).

## Data analysis

Statistical analyses were conducted using SPSS v.28 (IBM, 2021), supplemented by the PROCESS macro v.5.0 (Hayes, 2017). Alpha was set at.05. Prior to analysis, data were screened for accuracy and missing values. There were no missing responses in the main questionnaire scales, as the online survey design required items to be completed before submission. For demographic questions, participants were provided with an explicit ‘prefer not to answer’ option rather than leaving items blank, ensuring that missingness was not an issue in the dataset.

Preliminary analyses confirmed that the data met all assumptions for multiple regression. Assumptions of linearity, normality, and homoscedasticity were assessed through visual inspection of scatterplots and P-P plots (Tabachnick and Fidell, 2013). Independence of errors was confirmed via the Durbin-Watson test (= 1.91). Models were checked for multicollinearity, with all Variance Inflation Factors (VIF < 1.45) and Tolerance values (>m.69) falling well within acceptable limits. Finally, an analysis of Cook’s distance (max =.06) confirmed that no influential outliers exerted undue influence on the model results.

Hierarchical multiple regression was employed to examine the associations of service variables (i.e., rank, service years, voluntariness of exit, retirement length) in Step 1, perceived organisational support in Step 2, and resilience in Step 3 on retirement adjustment. To explore whether resilience moderated the relationship between perceived organisational support and retirement adjustment, a moderation analysis (Model 1) was conducted using the PROCESS macro. Although resilience may also function as a moderator, this role was examined on an exploratory basis.

Finally, to explore whether the association between organisational support and retirement adjustment varies over time, a second moderation analysis was performed with retirement length as a continuous moderator. The Johnson-Neyman (J-N) technique was utilised to identify the point in time at which the relationship between organisational support and retirement adjustment became statistically significant. In accordance with standard moderation procedures, continuous variables were mean-centred prior to analysis to produce more interpretable regression coefficients.

A mediation analysis (Model 4) was also performed to examine whether resilience accounts for the association between organisational support and retirement adjustment. This model evaluates statistical mediation based on cross-sectional covariance and does not provide definitive evidence of temporal or causal mediation. Bootstrapped samples (5,000, 95% CI) were calculated for the indirect effect. All continuous variables were mean-centred prior to analysis.

## Results

### Sample demographics

The final sample consisted of 289 retired officers, including those who retired on pensionable years, took early retirement, or retired on medical grounds. The mean age was 65.0 years (*SD* = 8.02, range 43-93), and there were 226 males (78.2%), 62 females (21.5%), and 1 participant who preferred not to report their sex (0.3%). The majority of respondents identified as ‘White English, Welsh, Scottish, Northern Irish or Irish’ (n = 280, 96.9%), and served in England or Wales (n = 272, 94.2%), with the remaining participants representing Scotland (n = 16, 5.5%) and Northern Ireland (n = 1, 0.3%). Most participants were married or in a civil partnership (n = 229, 79.2%), while others were divorced (n = 26, 9.0%), single (n = 10, 3.5%), or widowed (n = 10, 3.5%). The majority of the sample lived with a partner or spouse (n = 206, 71.3%) or with a partner/spouse and children (n = 35, 12.1%) or alone (n = 35, 12.1%). In addition, the majority indicated their departure was voluntary (n = 267, 92.4%), compared to 5.9% (n = 17) whose departure was involuntary (5 participants selected the option ‘prefer not to answer’). Participants had, on average, 25.86 years of police service (*SD* = 5.27, range 14–41) and had been retired for an average of 13.14 years (*SD* = 9.81, range 1 month – 50 years). Nearly half of the sample had served as Constables (n = 132, 45.7%), followed by Sergeants (n = 70, 24.2%). Inspector and Chief Inspector ranks were combined for reporting (n = 67, 23.2%), and Superintendent and Chief Superintendent ranks were similarly grouped (n = 19, 6.6%). One participant selected the option ‘prefer not to answer’(0.3%).

### Descriptive statistics and correlations

Descriptive statistics and bivariate correlations for the primary study variables are displayed in Table 1. On average, participants reported high levels of retirement adjustment (M = 46.31, SD = 9.61) and resilience (M = 3.36, SD =.70). The mean length of retirement for the sample was 13.14 years (SD = 9.81). Adjustment showed significant positive associations with all study variables, with the strongest correlation observed for resilience (r =.46, *p* <.001). In addition, we found a strong positive correlation between age and retirement length (r =.88, *p* <.001). Given this high degree of collinearity, retirement length was retained as the primary temporal moderator to specifically capture the transition period post-service, while acknowledging its role as a proxy for chronological age and generational cohort.

**Table 1.** Descriptive statistics and Pearson correlations between study variables.

| Variable | M | SD | 1 | 2 | 3 | 4 |
| --- | --- | --- | --- | --- | --- | --- |
| 1. Retirement adjustment | 46.31 | 9.61 | -- | .23*** | .46*** | .21*** |
| 2. Organisational support | 15.09 | 5.8 |  | -- | .32*** | .26*** |
| 3. Resilience | 3.36 | .70 |  |  | -- | .17* |
| 4. Retirement length | 13.14 | 9.81 |  |  |  | -- |

\* $p < .05$ , \*\* $p < .01$ , \*\*\* $p < .001$ .

### Predictors of retirement adjustment

A three-step hierarchical multiple regression was conducted with retirement adjustment as the outcome variable. In the first stage, retirement length and service years were entered, accounting for 4.5% of the variance [R^2^ =.05, Adjusted R^2^ =.04, *F*(2, 285) = 6.75, *p* =.001]. In the second step, rank and voluntary exit were entered, explaining an additional 6.0% of the variance [R^2^ =.10 Adjusted R^2^ =.08, *F*(4, 283) = 7.71, *p* <.001]. In the third step, perceived organisational support and resilience were entered. This final model was significant, *F*(6, 281) = 16.23, *p* <.001, and explained a total of 25.7% of the variance (R^2^ =.26, Adjusted R^2^ =.24). Notably, while retirement length (B =.22, β =.22, *p* <.001) and voluntary exit (B = - 6.95, β =-.19, *p* =.001) were significant predictors in Step 2, the effect of voluntary exit became non-significant (B =-3.57, β =-.10, *p* =.68) once resilience was included in Step 3. Resilience was the strongest predictor in the final model (B = 5.62, β =.41, *p* <.001), while perceived organisational support was not a significant predictor (B =.08, β =.05, *p* =.39). This suggests that resilience has a more direct association with retirement adjustment than organisational support in this model (see Table 2).

**Table 2.**
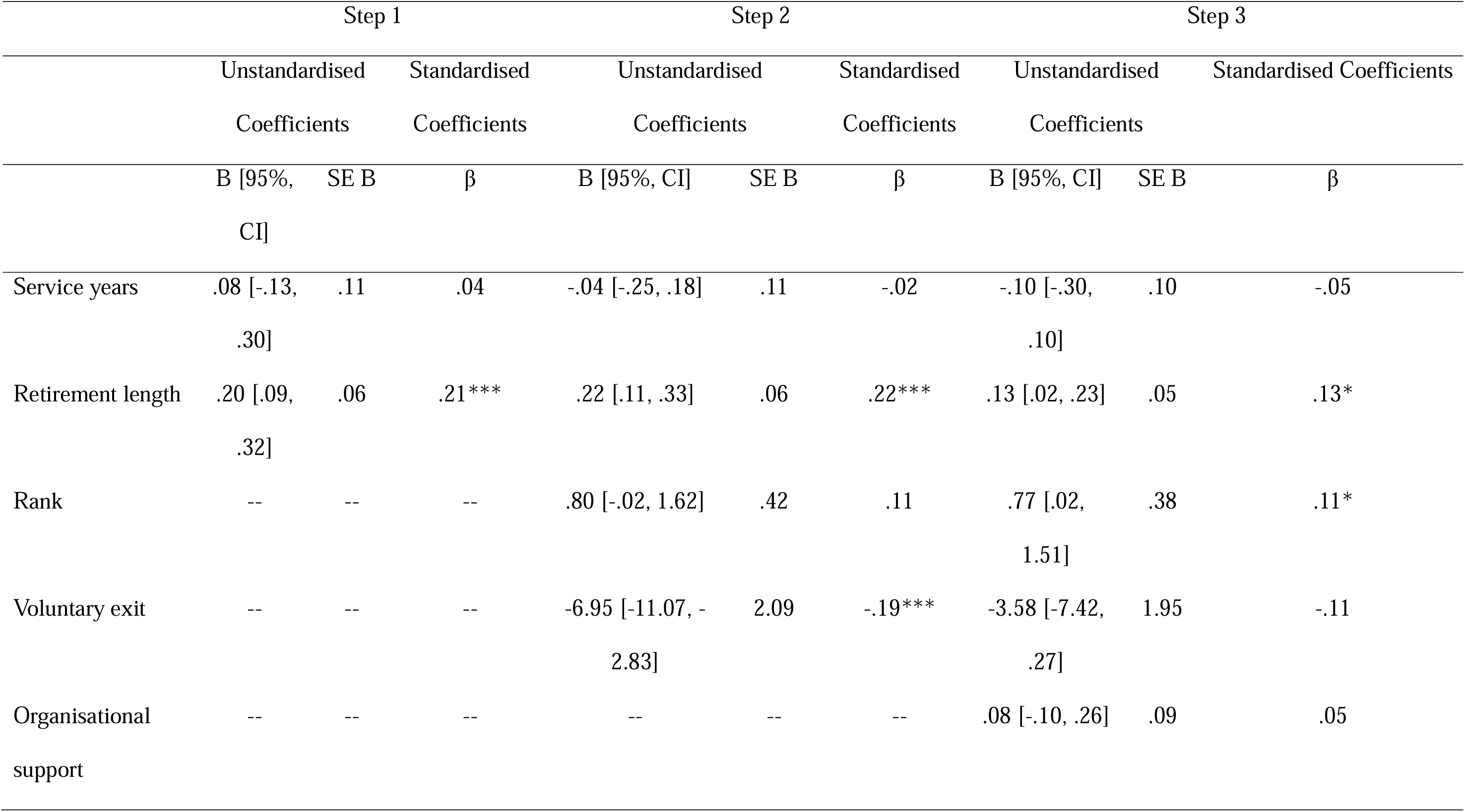
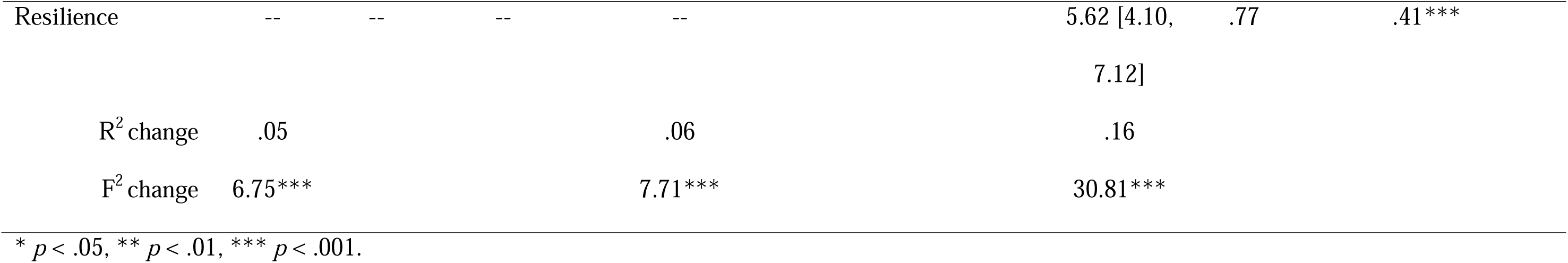
Hierarchical regression results predicting retirement adjustment.

To test whether resilience moderated the relationship between perceived organisational support and retirement adjustment, a moderation analysis was conducted. The overall model was significant, explaining 22.0% of the variance in retirement adjustment, *F*(3, 285) = 26.78, *p* <.001. Resilience was a significant positive predictor of adjustment (b = 6.01, *p* <.001). However, there was no significant main effect of perceived organisational support (b =.15, *p* =.112). The interaction was non-significant (b =.04 *p* =.71, ΔR^2^ <.001), indicating no evidence that resilience alters the strength of the association between organisational support and adjustment. These results suggest that resilience is independently associated with adjustment rather than functioning as a moderator of organisational factors.

### Temporal analysis of post-service adjustment

A moderation analysis was conducted to explore whether the association between organisational support and retirement adjustment varied by retirement length. The overall model was significant, explaining 24.6% of the variance in retirement adjustment, *F*(4, 283) = 23.06, *p* <.001. Resilience remained a significant independent predictor of adjustment (b =.88, *p* <.001). Although the interaction between perceived organisational support and retirement length was not significant and did not support the moderation hypothesis (b =.015, *p* =.065, ΔR^2^ =.01), an exploratory J-N analysis was conducted to examine whether conditional associations varied across retirement length.

J-N analysis suggested a trend whereby the association between organisational support and adjustment became stronger at longer retirement durations; however, this should be interpreted cautiously given the non-significant interaction (see Table 3). The relationship between organisational support and adjustment was significant only for those retired for more than 32.07 years (*p* =.05; see Table 4). Beyond this region of emergent salience, the strength of the relationship continued to increase as time since retirement progressed, reaching its largest observed association at the upper limit of the data (b =.50, *p* =.03).

**Table 3.** Exploratory Johnson–Neyman analysis of organisational support across retirement length.

| Retirement length (centred) | Raw years | b | SE | *p* |
| --- | --- | --- | --- | --- |
| -22.81 | 0 | -.27 | .22 | .22 |
| -10.31 | 12.5 | -.07 | .13 | .58 |
| -.31 | 22.5 | .08 | .09 | .39 |
| 9.14 | 32.07 | .23 | .11 | .05* |
| 14.69 | 37.5 | .31 | .15 | .03* |
| 27.19 | 50 | .50 | .24 | .03* |

\* $p < .05$ . SE = Standard Error

**Table 4.** Moderating effect of retirement length on the relationship between organisational support and adjustment.

| Outcome variable | Predictor variable | b | SE | *t* | *p* |
| --- | --- | --- | --- | --- | --- |
| Retirement adjustment |  | 26.26 | 2.59 | 10.12 | .001*** |
|  | Organisational support | .08 | .09 | .89 | .38 |
|  | Retirement length | .14 | .06 | 2.42 | .02* |
|  | Organisational support x Retirement length | .01 | .01 | 1.85 | .06 |
|  | Resilience (Covariate) | .98 | .13 | 7.74 | .001*** |

\* $p < .05$ , \*\* $p < .01$ , \*\*\* $p < .001$ .

Although these exploratory values give insight into potential temporal patterns, they do not provide evidence of a statistically supported moderation effect. Nevertheless, to visually illustrate this interaction, simple slopes were plotted at the 16th, 50th, and 84th percentiles of retirement length, representing 3.33, 13.14, and 23.95 raw years respectively (see Figure 1). As shown in the figure, the relationship between organisational support and adjustment was relatively flat for those in the early and mid-retirement phases (3.33-13.14 years). However, for those in the extended retirement phase (23.95 years), the slope was positive and steeper, illustrating the trend toward the region of emergent salience.

**Figure 1.** Interaction between perceived organisational support and retirement length on retirement adjustment. Higher scores reflect better retirement adjustment.

### Mediation analysis

To examine whether resilience accounts for the association between organisational support and retirement adjustment, a mediation analysis was conducted with resilience as the mediator. Results indicated a significant indirect effect linking organisational support to retirement adjustment through resilience (b =.23, Boot SE =.05). While the total effect of organisational support was significant (b =.38, *p* <.01), the direct effect became non-significant when resilience was included in the model (b =.15, *p* =.11). These findings are consistent with an indirect association whereby organisational support relates to adjustment through resilience.

## Discussion

The present study investigated the relationship between perceived organisational support, resilience, and retirement adjustment in retired UK police officers. By integrating Organisational Support Theory (Eisenberger et al., 1986) and Conservation of Resources theory (Hobfoll, 1989), this research sought to determine if individual resilience mediates the long-term impact of organisational experiences on retirement adjustment.

A primary finding of this study was that resilience accounted for the indirect association between perceived organisational support and retirement adjustment. This pattern provides empirical support for the integration of Organisational Support Theory (Eisenberger et al., 1986) and Conservation of Resources theory (Hobfoll, 1989). Instead of disappearing after exit, career-long contextual resources appear to align with enduring personal resource reservoirs that support post-service adjustment. In line with contemporary retirement adjustment frameworks (Wang et al., 2011; Wang and Huang, 2024), successful adaptation out of a high-stigma, high-demand uniform environment relies heavily on the successful capitalisation of these pre-retirement resource caravans. This mediation effect supports findings showing that pre-retirement resources are not static but serve as the foundation for adjustment and wellbeing post-retirement (Zhan et al., 2023). In this context, perceived organisational support functions as a contextual resource that strengthens individuals’ Pre□retirement reserves, thereby supporting a smoother transition into retirement (Wang and Huang, 2024).

In the high-stress environment of policing, organisational care is conceptually linked to the psychological capacity to bounce back from long-term exposure to trauma (Smith et al., 2008). When this support is present, officers transition into retirement with more robust resources, enabling better adjustment to retirement (Porter and Lee, 2024). On the contrary, a perceived breach in the reciprocal social exchange corresponds with lower psychological resource levels, reflecting higher vulnerability during transitions (Lennie et al., 2025; Farnese et al., 2018).

The temporal moderation analysis yielded a non-significant interaction term (*p* =.065), indicating that the hypothesis that retirement length moderates the relationship between organisational support and adjustment is not supported by the current data. Although an exploratory Johnson-Neyman analysis pointed toward a potential numerical trend indicating stronger associations at extended retirement durations, this pattern must be interpreted with caution. Given that the majority of police officers retire in their late 40s or 50s, reaching over three decades post-retirement places surviving retirees well into advanced old age, where well-documented post-service health attrition and longevity disparities significantly narrow the surviving cohort (Tanksley et al., 2025; Police Care UK, 2016). Consequently, rather than demonstrating a practical intervention target for middle-aged retirees, this late-emerging pattern should be viewed strictly as an exploratory cohort reflecting how a small subset of long-surviving officers retrospectively evaluate their careers.

Within these limits, the strength of the relationship continued to increase beyond this region of emergent salience as time since retirement progressed. This suggests that while resilience is a primary correlate of retirement adjustment throughout the post-service phase, the lingering legacy of the organisation may grow more salient as officers reach extended retirement. This late-blooming significance of organisational support contrasts with general longitudinal patterns where the impact of work-related factors typically diminishes as retirees move further from their career exit (Wang, 2007; Zhan et al., 2009). This may be due to a late-life re-evaluation of one’s career and sacrifice, a process aligned with lifespan developmental perspectives on life review and reminiscence in older age (Butler, 1963; Erikson, 1950). As individuals reach extended retirement and confront the final phases of the lifespan, they frequently engage in structured cognitive reviews of their past achievements and occupational identity to establish a coherent sense of meaning. Within this evaluative process, the retrospective belief that one’s former organisation valued their contribution (Eisenberger et al., 1986) serves as a vital narrative resource; recalling institutional validation helps mitigate feelings of bitterness regarding past sacrifices, thereby functioning as a psychological anchor that supports ongoing life satisfaction in late life.

This exploratory finding extends current understanding of the ‘cliff-edge’ of retirement by suggesting that aspects of the psychological contract (the reciprocal expectations between the officer and the force; Rousseau, 1989) may remain relevant decades after service (Lennie et al., 2025; Vaportzis & Edwards, 2026). To be precise, this’legacy’ effect is not demonstrated via intra-individual change over time, but is instead inferred from cross-sectional variation warranting future longitudinal verification. Accordingly, practical welfare recommendations should focus on proactive, lifelong support rather than waiting decades for late-stage retrospection.

Furthermore, this finding may reflect a generational shift in the policing experience. Older retirees served during an era characterised by a lack of formal welfare provision and support, where feeling valued was communicated through informal, direct supervisor support rather than institutional policy. For these officers, the personal nature of this support (or lack of it) creates a more persistent psychological legacy. As these retirees reflect on their service in older age, the perceived presence of this support acts as a durable resource that bolsters retirement adjustment, whereas its absence may lead to a retrospective sense of abandonment that hinders it.

### Theoretical and practical implications

In addition, the finding of resilience as a mediator addresses a critical gap in the literature regarding the specific pathways through which career-long experiences are translated into long-term outcomes (Birch, 2026). The role of resilience as a mediator addresses a critical gap in the literature by identifying it as the potential psychological pathway through which career-long organisational support leads to successful retirement adjustment. Finally, the identification of a region of emergent salience challenges the assumption that the impact of organisational factors diminishes linearly after exit, suggesting instead that the findings raise the possibility that perceptions of the psychological contract may remain relevant during later-life adjustment.

The findings also have practical implications for UK police policy and the Police Covenant (Home Office, 2024). If organisational support functions as a key resource associated with resilience, wellbeing cannot be viewed as a purely individual responsibility (Oscar Kilo, 2025). Police forces must recognise that the psychological implications of perceived organisational supportduring service may extend decades into retirement. Furthermore, interventions such as the RESLEAPS programme (a resilience-based training framework specifically designed for the police service) should continue to focus on bolstering individual psychological resources, as they are the primary mechanism for successful retirement adjustment (Lennie et al., 2025).

### Strengths, limitations, and future directions

A major strength is the inclusion of long-term retirees. By identifying the region of emergent salience, we provide exploratory evidence consistent with a possible persistent legacy of the psychological contract. In addition, we utilised Organisational Support Theory and Conservation of Resources theory to provide a robust explanation of both the mechanism (resilience) and the timing (the lingering legacy) through which work experiences impact retirees’ wellbeing. Furthermore, the findings provide direct empirical support for the UK Police Covenant, arguing that support is a lifelong commitment. These findings highlight the importance of embedding organisational support practices throughout an officer’s career, rather than concentrating resources solely at the point of retirement.

Despite these strengths, as a cross-sectional design was employed, causal inferences cannot be drawn from the observed associations. Consequently, the observed mediation pathways represent statistical mediation rather than true temporal mediation; longitudinal tracking is required to confirm causal pathways between organisational support, resilience, and adjustment. In addition, the sample may be subject to survivor bias; those with the poorest adjustment or worst health may not have been available to participate, suggesting the impact of poor support could be even more severe than reported. Future research should employ longitudinal methods to track the preservation of resources from the pre-retirement phase through several decades of post-service life. While offering valuable insights, our findings may not be generalisable to other demographic groups or international contexts as the sample was predominantly White British (96.9%) and male (78.2%). While this reflects broader historical demographics of UK policing, it limits the transferability of these findings; generational shifts and organisational cultures may manifest differently for female officers, ethnic minority groups, or international policing contexts where organisational cultures and welfare structures vary significantly. Research focusing on the retirement experiences of female and ethnic minority officers is essential to understand how different identities interact with organisational support. Furthermore, because pension eligibility ages and statutory retirement mandates differ globally, these findings should be generalised with caution across international jurisdictions.

Additionally, while the BRS demonstrated acceptable internal consistency (α =.71), this modest reliability warrants cautious interpretation. The measurement of resilience remains subject to ongoing academic debate regarding whether it functions best as a stable baseline trait or as a multi-systemic, dynamic process of adapting to adversity over time (Denckla et al., 2020). Future research should employ multi-dimensional or longitudinal assessments of resilience to capture how these fluctuating coping processes evolve across the post-service lifespan. Finally, the data rely on the participants’ memories of how they were treated decades ago. While this perceived support is what matters for their current mental state, it may be subject to recall bias, although perceived support remains the most relevant factor for current psychological states.

## Conclusion

The findings provide empirical evidence of a statistically significant indirect association between perceived organisational support and retirement adjustment through resilience, confirming resilience as an important mediator. Within the framework of Conservation of Resources theory, these results suggest that organisational support functions as a vital contextual resource associated with the personal psychological resources necessary for a successful transition into post-service life (Hobfoll, 2012; Wang et al., 2011). While resilience remains the most robust independent predictor of adjustment, the influence of the organisation does not diminish upon exit. Regarding the temporal moderation model, the overall interaction term was non-significant; however, exploratory Johnson-Neyman analysis identified a potential numerical trend indicating that retrospective perceptions of organisational value grow more salient beyond 32 years of retirement. Although this late-emerging legacy pattern must be interpreted with caution, it highlights how a subset of long-surviving officers retrospectively evaluate their careers. Ultimately, fostering an organisational culture of support is not merely an in-service requirement but a long-term investment in the successful life adjustment of those who have served.

## Ethical approval and informed consent statements

Ethical approval was granted by the Humanities, Social, and Health Sciences Research Ethics Panel at the University of Bradford (Reference Number E1350). All participants provided informed consent prior to participation. **Data availability** The datasets generated during and/or analysed during the current study are available from the corresponding author on reasonable request.

## Author contributions

*Eleftheria Vaportzis*: Conceptualization, Methodology, Formal Analysis, Writing-Original draft preparation, Writing-Reviewing and Editing, Project Administration.

*Warren Edwards*: Conceptualization, Formal Analysis, Writing-Reviewing and Editing.

## Data Availability

All data produced in the present study are available upon reasonable request to the authors

